# Pain and cognitive performance in adults with multiple sclerosis: A systematic review

**DOI:** 10.1101/2022.12.09.22283279

**Authors:** Fraser S Brown, Stella A Glasmacher, Daniel Taylor, Ruth Jenkins, Siddharthan Chandran, David Gillespie, Peter Foley

## Abstract

**Introduction:** Pain and cognitive dysfunction are separately known to be important manifestations of multiple sclerosis (MS). Although pain is a complex subjective phenomenon with affective and cognitive aspects, it is not known if people with MS reporting pain are at greater risk of reduced performance in objective tests of cognition. The presence or direction of any association remains to be clarified, as do the roles of confounders such as fatigue, medication and mood.

**Methods:** We conducted a systematic review of studies examining the relationship between pain and objectively measured cognition in adults with confirmed MS, according to a pre-registered protocol (PROSPERO 42020171469). We carried out searches in MEDLINE, Embase and PsychInfo. We evaluated the role of potential confounders (medication, depression, anxiety, fatigue and sleep) and described findings by eight pre-specified cognitive domains.

**Results:** 11 studies (n=3714, range 16 to 1890 participants per study) were included in the review. Four studies included longitudinal data. Nine studies identified a relationship between pain and objectively measured cognitive performance. In seven of these studies, higher pain scores were associated with poorer cognitive performance. However, no evidence was available for some cognitive domains. Heterogeneous study methodology precluded meta-analysis. Studies infrequently controlled for the specified confounders. Most studies were judged to be at risk of bias.

**Discussion:** Several studies, but not all, identified a negative relationship between pain severity and objectively measured cognitive performance. Our ability to further characterise this relationship is limited by study design and lack of evidence in many cognitive domains. Future studies should better establish this relationship and delineate the neurological substrate underpinning it.

## Introduction

Multiple sclerosis (MS) is an autoimmune and neurodegenerative disorder of the central nervous system. Demyelination and neurodegeneration are both recognized and are key substrates of relapsing remitting or progressive subtypes, respectively.(1) Presentation with visual, motor or sensory abnormalities is considered typical but co-existing or prodromal ‘invisible’ symptoms such as cognitive dysfunction, pain and fatigue are increasingly recognised.(2) Priority-setting groups comprised of patients, carers and clinicians have identified pain and cognition as priority areas in MS research.(3) Such symptoms have a significant impact on quality of life and may have greater impact than those symptoms traditionally thought to be disabling.(1, 4)

Cognitive impairment is extremely common in MS, with prevalence up to 70% (5) After initial recognition by Charcot and relative obscurity during much of the 20^th^ century, cognitive dysfunction is now an increasingly researched problem in MS.(6) Multiple cognitive domains can be affected, with processing speed and episodic memory reported to be those most frequently impaired.(6-9) Executive function, visuospatial and verbal fluency are also often impaired.(6-9) Cognitive impairment appears to be more common in progressive than non-progressive disease.(10)

Pain is also common in MS, with estimates of prevalence ranging between 29 and 86%.(4,11,12) Like cognitive dysfunction, pain may be more common in progressive MS.(13) Various pain syndromes, including neuropathic pain, headaches (including trigeminal neuralgia), back pain and spasms are common(12). However, the underpinning pathophysiological mechanisms remain elusive. (12) The evidence base to guide pharmacological treatment of pain in MS is limited (14,15) with clinical guidelines extrapolating evidence from studies in other clinical conditions.(16) Similarly, there is a paucity of evidence for non-pharmacological treatments for pain in MS.(17). There is limited evidence regarding imaging correlates of pain in MS.(18,19)

Chronic pain and cognitive dysfunction are intimately linked (20). Multiple cognitive processes contribute to the experience of pain, in particular executive function (20). Indeed, nociception is heavily reliant on cognitive processes, which may lead to disparity between insult and the subjective experience and reporting of pain (21). The neural substrate underpinning these observations is beginning to be probed by functional imaging studies; distraction from pain has been shown to attenuate imaging correlates of pain (21). Coined ‘attentional analgesia’, this phenomenon may be underpinned by bidirectional cortex-brainstem neural pathways (22). As such, aberrant cognitive processing, a moderating step in the nociceptive pathway, may influence perceived pain.

Conversely, pain might also influence cognitive processing (23). Data from patients with Alzheimer’s disease support this observation. Whitlock et al showed higher rates of subsequent cognitive decline in patients with persistent chronic pain.(24) They propose that pain may subvert cognitive processing resources and therefore exacerbate cognitive decline over time. As such, one proposed relationship is that pain may precede cognitive dysfunction. An alternative hypothesis may be that pain and cognitive dysfunction are both independently associated with more severe illness. Furthermore, many analgesics may impact cognition (for example, tricyclic antidepressants)(25) underscoring the importance of identifying any association of pain and cognition in adults with MS.

Ultimately, improved understanding of the relationship between pain and cognitive performance may better direct research goals and therapeutic efforts. Specifically, it remains unclear if pain in people with MS is associated with poorer cognitive performance, or increased risk of future cognitive dysfunction. Clarification of this relationship could, for example, guide focused examination of cognition when pain is identified, or assist in development of psychological therapies for pain. Here, we conduct a systematic review of prospective studies to determine the relationship between presence and severity of pain, and objectively measured cognitive performance, in adults with multiple sclerosis.

## Methods

This work adheres to the Preferred Reporting Items for Systematic Reviews and Meta-analyses standards (PRISMA).(26) We conducted this review according to a pre-specified protocol registered with PROSPERO (CRD42020171469). We searched MEDLINE, Embase and PsycINFO between inception and 12^th^ April 2022 (see supplemental file for full search strategy). Peer-reviewed original research was included. There were no language restrictions. Two authors independently reviewed titles, abstracts and full text for inclusion; disagreements were resolved by review of third author or discussion.

We included studies of adults (age ≥18 years old) with confirmed MS (any subtype) diagnosed by a physician, which reported assessments of pain and objective cognitive performance using validated instruments. Cohort, cross-sectional and randomized controlled trial designs were included (in the latter, we analysed baseline data prior to intervention). We included studies measuring objective rather than participant-reported subjective cognitive performance as the latter may be more often confounded by mood and depression.(27) We did not require use of specific contemporaneous diagnostic criteria for multiple sclerosis, though these were recorded where presented. Exclusion criteria included non-human studies, paediatric populations (age <18), articles other than original research, pain due to a specific intervention and non-validated cognitive function measures. Cognitive assessment instrument validity and modality was determined by a neuropsychologist with experience of working with people with MS (DG). Each neuropsychological test was assigned to one of eight pre-specified cognitive domains according to an internationally recognized classification and in line with previous work(28, 29), namely: orientation and attention, language, concept formation and reasoning, executive function, memory, construction and motor praxis, perception and global cognition. The single digit modality test (SDMT), which measures processing speed, is categorized under orientation and attention.(28)

Data extraction was performed by two authors using a pre-specified tool. A pre-determined modified Newcastle-Ottowa scale (NOS)(30) was used to assess study quality. Each record was analysed by two authors independently. Study quality was scored on factors judged by the review authors to be important in the assessment of the pertinent variables of pain and cognition. Points were awarded for cohort representativeness (lack of selection bias), sample size >50, description of MS subtype, description of educational attainment, control for depression or fatigue, control for medication, longitudinal design, blinded assessment of outcome and statistical data presented. The agreement between authors was quantified using Kohen’s kappa. We defined very high, high and low risk of bias as 0-3, 4-6 and 7-9 NOS points, respectively, in line with previous work.(31)

## Results

### Search results, study design and participants

The search strategy yielded 1790 studies overall of which 11 were included (Following PRISMA guidelines; see Figure 1).(32-42)

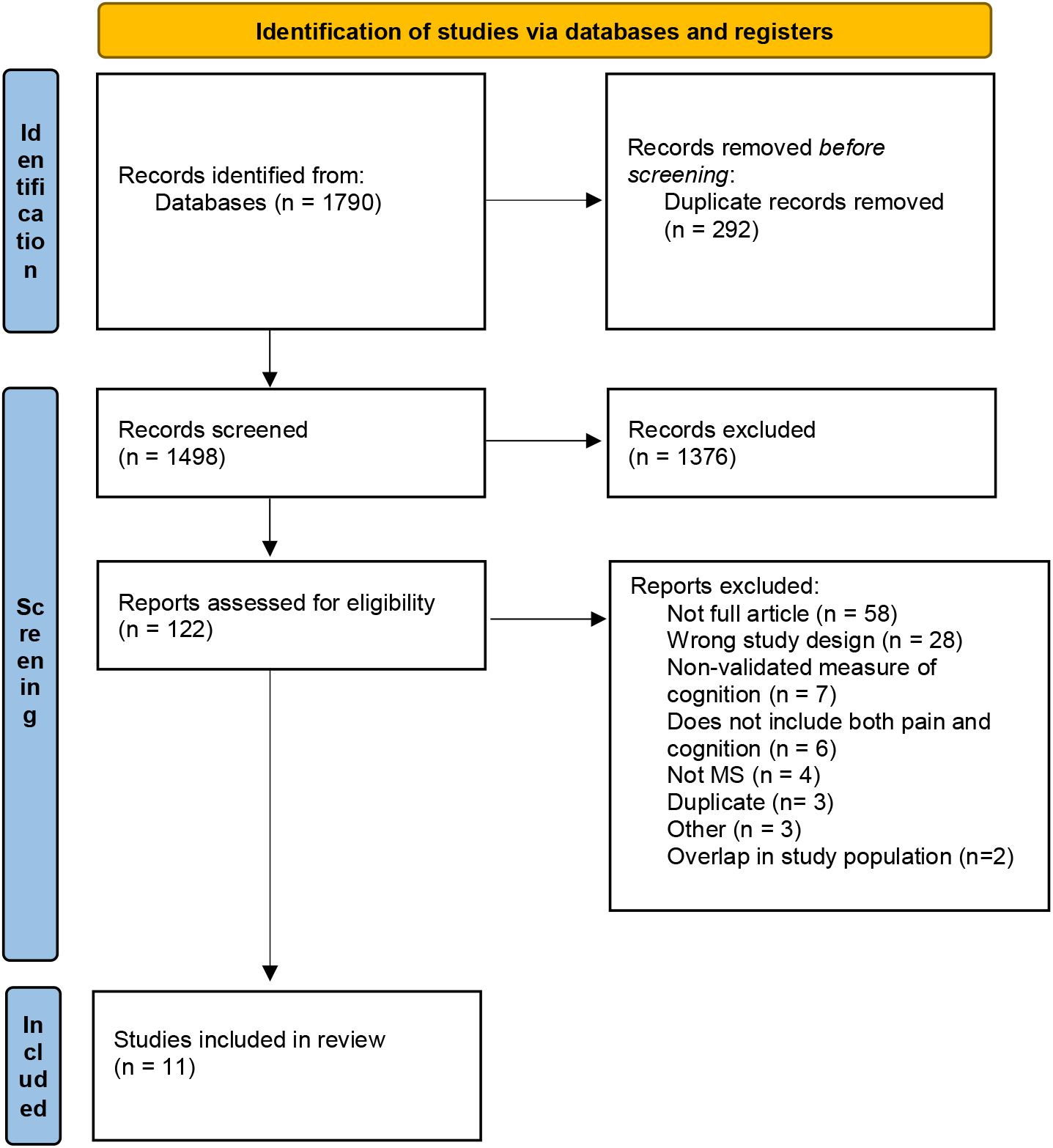
PRISMA 2020 flow diagram for new systematic reviews which included searches of databases and registers only. *From:* Page MJ, McKenzie JE, Bossuyt PM, Boutron I, Hoffmann TC, Mulrow CD, et al. The PRISMA 2020 statement: an updated guideline for reporting systematic reviews. BMJ 2021;372:n71. doi: 10.1136/bmj.n71 For more information, visit: http://www.prisma-statement.org/

Study designs of included studies were cross-sectional (n=7)(34, 36, 38-42) cohort (n=3) (33, 35, 37) and randomized controlled trial (n=1)(32). Sample size ranged from 16 to 1890 people with MS (mean 338). The proportion of female patients in studies ranged from 45.8% to 100%. Mean participant age ranged from 34 to 59.6 years. Nine studies were based in the community (32, 33, 35-37, 39-42) and two in a long-term care home setting (34, 38). Educational attainment was recorded in eight studies.(33, 35-40, 41) Studies were carried out in the United States,(33, 34, 36-39) France,(32) Germany,(35) the Netherlands(40, 41) and Australia (42)

Studies included participants experiencing relapsing remitting, secondary progressive, and primary progressive MS subtypes. One study included participants classified as both MS and clinically isolated syndrome (CIS), as a single group.(35)

### Pain and cognition measurements

The type of pain being examined was poorly reported across studies. Two studies specified neuropathic pain.(32, 35) Pain type was unspecified in the remaining studies.(33, 34, 36-42) In ten studies, pain severity was measured.(32-36, 38-42). Pain presence as a binary variable was measured in one study.(37) A range of neuropsychological assessments were used to measure cognitive performance (Table). Studies reported assessments of the following cognitive domains: orientation and attention (n=8 studies) (32, 34, 35, 38-42) memory (n=3 studies) (36, 40, 41), global cognition (n=4 studies) (33, 37, 40, 41) and executive function (n=2 studies) (40,41). No studies employed measures of language, concept formation and reasoning, construction/motor praxis, or perception. Therefore evidence was available for only four of the eight (28) pre-specified cognitive domains.

**Table 1.** Summary of association between pain and cognition.

|  |  |  | **Lezak category**<br>(where applicable, specific cognitive function tested is detailed) |  |  |  |  |  |  |  |
| --- | --- | --- | --- | --- | --- | --- | --- | --- | --- | --- |
| **Paper** | **Severity or presence of pain** | **Cognitive assessment** | Orientation and attention | Language | Concept formation and reasoning | Executive function | Memory | Construction and motor praxis | Perception | Global cognition |
| Ayache 2016 | Severity | ANT |  |  |  |  |  |  |  |  |
| Demakis 2010 | Severity | MDS-cog for nursing home residents |  |  |  |  |  |  |  |  |
| Fritz 2016 | Severity | SDMT |  |  |  |  |  |  |  |  |
| Heitmann 2020 | Severity | PASAT | Attention and processing speed |  |  |  |  |  |  |  |
| Miller 2014 | Severity | MIST and its subscales |  |  |  |  |  |  |  |  |
| Newlands 2005 | Presence | Cognitive performance scale and its subscales |  |  |  |  |  |  |  |  |
| Newlands 2012 | Severity | PASAT | Attention and processing speed |  |  |  |  |  |  |  |
| Sandroff 2017 | Severity | SDMT | Processing speed |  |  |  |  |  |  |  |
| Scherder 2017 | Severity | MMSE |  |  |  |  |  |  |  |  |
|  |  | Eight words test |  |  |  |  | Verbal memory |  |  |  |
|  |  | Face recognition, (Rivermead) |  |  |  |  | Visual memory |  |  |  |
|  |  | Picture recognition, (Rivermead) |  |  |  |  | Visual memory |  |  |  |
|  |  | Category fluency (a subtest of the Dutch Groninger Intelligence Test) |  |  |  | Visual fluency |  |  |  |  |
|  |  | Digit span backward, (Wechsler) | Orientation and memory |  |  |  |  |  |  |  |
|  |  | Key search (BADs) |  |  |  | Planning |  |  |  |  |
|  |  | Rule shift cards (BADs) |  |  |  | Cognitive flexibility |  |  |  |  |
| Scherder 2021 | Severity | MMSE |  |  |  |  |  |  |  |  |
|  |  | Eight words test |  |  |  |  | Verbal memory |  |  |  |
|  |  | Face recognition, (Rivermead) |  |  |  |  | Visual memory |  |  |  |
|  |  | Picture recognition, (Rivermead) |  |  |  |  | Visual memory |  |  |  |
|  |  | Digit span backward, (Wechsler) | Orientation and memory |  |  |  |  |  |  |  |
|  |  | Rule shift cards (BADs) |  |  |  | Cognitive flexibility |  |  |  |  |
|  |  | Category Fluency (a subtest of the Dutch Groninger Intelligence Test) |  |  |  | Visual fluency |  |  |  |  |
| Spain 2007 | Severity | SDMT | Information processing speed |  |  |  |  |  |  |  |

no relationship identified
worse pain associated with better cognitive performance
worse pain associated with worse cognitive performance
Abbreviations: ANT, Attention Network Test; MDS-cog, Minimum Data Set Cognition Scale; SDMT, Single Digit Modalities Test; PASAT, Paced Auditory Serial Addition Test; MIST, Memory for Intentions Test; MMSE, Mini Mental State Examination; BADs, Behavioural Assessment of the Dysexecutive Syndrome.

### Relationship between pain and cognition

Statistical methods used to examine the relationship between pain and cognition included linear regression,(35, 36, 40, 41) logistic regression,(37, 41) Pearson correlation,(39) T Test,(32) Spearman’s correlation,(34) principal component analysis(38) and hierarchical regression analysis.(33)

Nine studies identified a relationship between pain and cognition.(33-39, 41, 42) Eight of these studies reported statistically significant association by varying methods; in the ninth study, the symptoms were associated in a symptom cluster by principal component analysis.(37)

In seven studies, higher pain scores were negatively associated with objective cognitive performance.(34-36, 38, 39, 41, 42) In all of these studies, pain severity was the variable measured. Three of these studies measured processing speed using SDMT.(43) Two measured attention and processing speed using Paced Auditory Serial Addition Test.(44) One measured orientation and memory using the digit span backwards test.(45) The final study measured memory using the memory for intentions test.(46)

In two studies, worse pain was associated with better cognitive performance, including in longitudinal analyses.(33, 37) In contrast to other included studies, both of these studies employed measures of global cognition in nursing home residents, the Minimum Data Set Cognition Scale (MDS-Cog) and the Cognitive Performance Scale (CPS), respectively.(47, 48).

Four studies (32, 33, 35, 37) collected longitudinal data, including the two studies of nursing home residents employing MDS-Cog and CPS (33, 37). Longitudinal studies were interpreted by the authors as showing: no association between pain and cognition (32); worse pain predicted better cognition at a one year interval(33); a cross-sectional association at baseline without longitudinal effect of pain on later cognition (higher pain score negatively associated with cognitive performance at baseline, cognition at baseline did not predict pain at one year) (35) and baseline pain predicted better cognition at a 180 day interval (37).

### Quality Assessment

Quality assessment of each study by two independent authors showed high agreement (Kohen’s kappa = 0.8). Given high agreement, results from one reviewer who assessed all studies were used for further analysis. Generalisability of the cohorts was felt to be limited in n=8 studies.(32, 34, 36, 38-42) Sample size was greater than 50 people in n=7 studies.(33, 35-37, 39-41), MS subtype was specified in n=8 publications.(32, 34-36, 38, 39, 41, 42) Educational attainment was described in eight publications(33, 35-40, 41) but controlled for only in three.(33, 35, 36) Depression was controlled for in five studies(33, 35, 36, 40, 41) while no studies controlled for anxiety. Fatigue was controlled for in one study.(35) No studies controlled for medication use or sleep disturbance. Only one study described blinding of investigators.(40) Statistical analysis was presented in all papers studied. We were unable to undertake meta-analysis due to heterogeneity in study design.

NOS quality assessment yielded overall scores of between two and seven. Two studies were rated as very high risk of bias (NOS score 0-3),(34,38) eight at high risk (4-6)(32, 33, 36, 37, 39-42) and only one at low risk of bias (NOS score 7-9)(35).

Notably, the one study judged to be at low risk of bias reported that higher levels of pain were associated with poorer objective cognitive performance.

## Discussion

Here, we present evidence for an association between cognition and pain in MS based on systematic review. We identified 11 studies, including over three thousand adult participants with MS, that allowed examination of the relationship between pain and objective cognitive performance. Although most studies included identified that higher levels of pain were associated with worse cognitive outcomes, two studies reported that worse pain was associated with better cognitive outcomes. (33, 37) Both of these studies were carried out in nursing home residents using observer-administered global cognitive assessments-MDS-COG and CPS, respectively.(47, 48) Our ability to further quantify the relationship between pain and cognition in adults with MS is limited by heterogeneity in study design (precluding meta-analysis), and limitation in the neuropsychological domains assessed in the existing literature.

We observed limitations in the identified evidence. The cross-sectional nature of the included studies limits conclusions regarding causality. Future studies using longitudinal designs will allow optimal assessment of the relationship between pain and performance on cognitive testing. Additionally, multiple factors could mediate this relationship (24). These include concomitant medicines and comorbidities. In the included studies, some confounders were rarely controlled for. Educational attainment is one such important factor. Some studies did account for educational attainment but often this was a relatively crude delineation eg. high school completion, and may not adequately account for this important confounder.(37) Three studies did not report any measure of educational attainment. Medication was not controlled for in any study, despite the potential for some classes of analgesic medicines to affect cognitive performance.(25) These unmeasured factors may therefore confound study conclusions. Controlling for confounders will be essential in future work. Additionally, most studies did not describe blinding of investigators, a potential source of bias.

Further, the patient populations included were heterogeneous regarding disease subtype. Therefore, generalisability of these data to adults experiencing other MS disease subytpes should be a focus of future study. Importantly, given that cognitive deficits may be more prevalent in progressive disease (10), a preponderance of a relapsing remitting phenotype in some studies may reduce applicability of the findings to the population with the highest burden of cognitive dysfunction. It is also notable that most patients included here were recruited from community settings, potentially limiting generalizability to inpatient or long-term care facility settings. Furthermore, the studies were performed in the United States, Europe and Australia which may limit applicability to cohorts in other countries given recognized geographical variation in disease course.(49)

A heterogeneous range of cognitive assessments was employed, probing different cognitive domains (orientation and attention, memory, executive function and global cognition). Pain may impact some cognitive domains more than others. We cannot exclude the possibility of relationships between pain and specific cognitive domains where evidence is not available. Specifically, further study of the relationship between pain and executive function will be particularly important. The nature of the pain syndrome experienced was also poorly reported in the included studies. Notably, while many of the included studies did not specify duration of pain it is likely that most participants were reporting chronic pain issues in the observed period. Future work is required to better define the relationship between pain and cognition by studying more defined pain syndromes. In some studies the relationship between pain and cognition was not the primary research focus; therefore study methodology may not have been optimally designed for investigating these variables. Additionally, publication bias, wherein negative results are less likely to have been published, may also affect our results.

Two studies identified that pain was associated with better scores in cognitive assessment (33,37). In these studies of nursing home residents, observer-applied assessment methods for pain and cognition were used.(33, 37) These methods relied on ability to communicate - indeed, communication ability formed part of the cognitive assessment. Pain assessment was based on frequency of observed or subject-communicated pain. In these studies specifically, a better cognitive score (partly based on ability to communicate) could be associated with ability to *communicate* pain (and therefore a higher pain score). More cognitively impaired patients, for example those with dementia, may have very limited communication, leading to spuriously low reporting of pain in this group. For these reasons, we suggest that data identifying a link between patient-reported pain, and cognitive performance in these studies, should be interpreted with caution.

Interestingly, these two studies did not include attentional cognitive tests which were widely used in other included studies but rather focused on memory, orientation, decision making, self-expression and tasks such as dressing.(33, 37) Another possible explanation is that pain exerts a cognitive modality specific effect. As such, it could be possible that attention-focused cognitive dysfunction is more susceptible to pain-mediated interference. Previous findings support a domain-specific effect as we have alluded to here.(50-52)

Strengths of our study include attribution of tested cognitive functions to pre-specified categories supported by specialist neuropsychology input. This approach allowed assessment of relationships between pain and individual cognitive domains. We included only objective measures of cognition, which should act to limit confounding by mood.(27) Subjective estimates of cognitive ability may be particularly closely related to affective disturbances. Indeed, subjective difficulty in concentrating is a core diagnostic criterion for diagnosis of depression according to DSM-5 criteria(53) and is included in research instruments measuring depressive symptoms (54). The present study included all subtypes of MS and accepted historical diagnostic criteria; while this approach was chosen to maximize sensitivity, it does have attendant caveats.

One such limitation may be the inclusion of a cohort where a proportion of patients had CIS (35) This cohort was included given both the likelihood some of those with CIS would be diagnosed with MS under contemporary diagnostic criteria, and that many went on to develop MS. 88% of CIS patients in this study met the authors’ criteria for a formal diagnosis of RRMS by the end of the study period.(35) This highlights the potential heterogeneity of study participants over time given evolution of MS diagnostic criteria and inherent limitations in applying historical studies to modern practice.

Additionally, cautious interpretation is required regarding NOS scores. There is no consensus on thresholds distinguishing quality categories. We chose to define categories here based on previous work in lieu of established formal cut-offs.(31) Even accounting for interpretation of NOS score categorization, the majority of studies included appeared to be at risk of bias.

## Conclusion

Here, we present a systematic review examining the relationship between cognition and pain in MS. Seven of eleven studies identified that higher levels of pain were associated with poorer cognitive performance. Two studies identified that higher levels of pain were associated with better cognitive performance, however interpretation of these findings is limited by study methodology. The identified evidence suggests that pain and cognitive performance could be inter-related in people with MS. Studies included were however heterogeneous, and important confounders were infrequently studied. So-called invisible symptoms of MS remain relatively undertreated; improved understanding of relationships between pain and cognition in MS, in particular any causal relationships, might lead to improved therapeutic strategies. Future work should attempt to elucidate this interaction and its neural substrate(s), as well as explore therapeutic effects on these symptoms.

## Supporting information

Search Strategy

PRISMA checklist

## Data Availability

All data described in the present work are contained in the manuscript

## Notes

### Competing Interest Statement

The authors have declared no competing interest.

### Funding Statement

This study received no specific funding

