## Supplementary material for "Pain and cognitive performance in adults with multiple sclerosis: A systematic review": Search Strategy

Ovid PsycINFO:

1. multiple sclerosis/
2. exp Pain/
3. neuropathic pain/
4. 2 or 3
5. cognitive impairment/
6. cogniti*.mp.
7. 5 or 6
8. 1 and 4 and 7

Ovid MEDLINE(R):

1. Multiple sclerosis/
2. exp Pain/
3. exp neuromuscular manifestations/
4. Cognitive dysfunction/
5. Cogniti*.mp
6. communication disorders/ or confusion/ or memory disorders/ or intellectual disability/
7. 2 or 3
8. 4 or 5 or 6
9. 1 and 7 and 8

Ovid EMBASE:

1. Multiple sclerosis/
2. Pain/
3. Chronic pain/
4. Chronic inflammatory pain/
5. Face pain/
6. exp “headache and facial pain”/
7. exp leg pain/
8. exp limb pain/
9. neck pain/
10. neuropathic pain/
11. cognitive defect/
12. cogniti*.mp
13. 2 or 3 or 4 or 5 or 6 or 7 or 8 or 9 or 10
14. 11 or 12
15. 1 and 13 and 14
